# P1 variants and key amino acid mutations at the Spike gene identified using Sanger protocols

**DOI:** 10.1101/2021.03.21.21253158

**Authors:** Gabriela Bastos Cabral, Cintia Mayumi Ahagon, Giselle Ibette Silva Lopez-Lopes, Igor Mohamed Hussein, Paula Morena Guimaraes, Audrey Cilli, Valeria Oliveira Silva, Maria do Carmo CT Timenetsky, Ivy de Jesus Alves, Andrea GC Bombonatte, Fabiana C Pereira dos Santos, Luís Fernando de Macedo Brígido

## Abstract

SARS-CoV-2 variants, along with vaccination, mark the second year of the pandemic. The spike region is a focal point in COVID-19 pathogenesis, with different amino acid changes potentially modulating vaccine response and some being part of variant signatures. NGS is the standard tool to sequence the virus but limitations of different sources hinders expansion of genomic surveillance in many places. To improve surveillance capability we developed a Sanger based sequencing protocol to obtain coverage of most (>95%) spike gene. Eleven nasopharyngeal swabs collections had RNA extracted for real time PCR diagnosis and leftover RNA had up to 3785 bp sequenced at an ABI3500 using dye termination chemistry of nested PCR products of two reactions of one-step RT-PCR. P1 amino acid mutations signatures were present in 18% (2/11), with 82% (9/11) with three or more additional amino acid changes (GISAID CoVsurver list). Most sequences (86%, 6/7) from 2021 have the E484K, whereas the mutation was not present in samples collected in 2020 (0/4, p=0.015).The swiftness that favorable mutations to the virus may prevail and their potential impact in vaccines and other current interventions need broader surveillance and more public health attention.

## INTRODUCTION

As COVID-19 pandemic enters its second year, positive signs from advance vaccination coverage brings hope amid an unsettled scenario of new infection waves in many areas. New variants have been increasingly detected, and have been associated to increased Infectivity (Korber, 2020), transmissibility (Kirby, 2021) and reinfection (Naveca, 2021). Severity of disease is more difficult to access and data is not conclusive, but impact of variants in vaccine response is key to pandemic control. Signs of concern have emerged both from in vitro neutralization studies from convalescent plasma (Liu, 2021; Wilfredo, 2021) and post vaccination plasma (de Souza, 2021) as well as evidence from vaccine trials (Shabir, 2021). In this setting molecular epidemiology assumes an even more important role not only for monitoring the evolution of the virus but also to inform on potential impact on plasma or monoclonal antibodies therapies as well as in vaccine strategies.

Next generation sequencing (NGS) is the standard technique to study SARS-CoV-2 genome variability. However, the worldwide capacity is limited, the technique demands specific requirements and it is not available in many areas. Sanger protocol, central to molecular epidemiology of HIV and other agents before the NGS area, may provide important information that may complement NGS, as screening samples for further NGS analysis.

New variants and vaccines have in common the Spike region, a 1292 amino acid protein that includes the receptor-binding domain (RBD). It is the active immunogen of most vaccines products and harbors many of the mutations described in the most known new variants, as the UK variant (B1.1.17), the South Africa (B.1.351) and the Brazilian Amazon (B.1.1.28, P1) variants. These variants, albeit showing mutations in other regions of the SARS-CoV-2 genome, have many amino acid substitutions or deletions at the Spike gene. One of the key mutations in these variants is the N501Y amino acid change at RDB, that increase biochemical binding capability to the mutated protein (Haolin & Liu, 2021). Another key mutation is E484K, an amino acid change that has been shown in vitro to decrease ability of plasma from vaccinated individuals to block viral entry (Collier, 2021).

The identification of mutation signatures for these new variants may serve as a proxy for the presence of a variant in the population studied, that eventually can be further evaluated by NGS. Therefore, the Spike gene itself seems a reasonable target to monitor the variants already identified and new emerging mutations that may give rise to new variants. Moreover, Spike region analysis of cases of infections after vaccination will be instrumental to monitor vaccine effectiveness as the evolution of new variants of the virus.

To improve genomic monitoring capability, we developed a simple protocol using one-step PCR to amplify DNA sequenced using classical “BigDye/Sanger” platforms.

## PATIENTS AND METHODS

RNA extracted from 11 nasopharyngeal swab (SWNF) samples were used to conduct the molecular assays, processed at the Virology Center or at the Regional Adolfo Lutz Center. All samples were previously tested by RT-qPCR (CDC, 2020) that confirmed for SARS-CoV-2 infection.

### Primers design

In order to design primer suitable PCR sets to partial SARS-CoV-2 Spike (S1 and S2 region) protein, 3 sequences (NC_045512.2); SARS-CoV-2 Wuhan-Hu-1, and two 2 sequences from the first case reported in Brazil, MT350282.1 and MT126808.1; (Araujo, 2020) were obtained from NCBI and imported into BioEdit sequence alignment editor (version 7.0.5.2) program.

The process of primer designing was conducted manually, and no automated software packages used. The primers were design to conduct a nested reverse transcription-polymerase chain reaction (RT-PCR) protocol.

This protocol allows the amplification of up to 3785 nucleotides of the Spike protein, using two one-step PCR and 4 semi-nested-PCR. A fragment comprising the beginning of the S1 (complete region 253/253a.a) to partial S2 (561/609 a.a) and another for the S2 region (609a.a/1827pb) corresponding to 2586 pb of Spike protein.

For Spike S1 protein amplification the primers set used were: (i) First round (one-step RT-PCR); COV_1out_Forward AGG GGT ACT GCT GTT ATG TC (21421-21440) and COV_2_out_Reverse GCA CCA ATG GGT ATG TCA CA (23545-23565), resulting in a 2,144 kb product; and (ii) Second round (nested PCR) COV_1out_Forward AGG GGT ACT GCT GTT ATG TC (21421-21440) and COV_3_Inner_Reverse ATCAGCAATCTTTCCAGTTTGC (22801-22823pb), generating a fragment of 1,402 Kb (S1-A). COV_4out_Forward AGTGTTATGGAGTGTCTCCTACT (22695-22718) COV_2_out_Reverse GCA CCA ATG GGT ATG TCA CA (23545-23565) generating a fragment of 0,870 Kb (S1-B). Primers pairs allowed the amplification of S1 protein, comprising nucleotide position 22549-23310 (253 a.a) based on SARS-CoV-2 Wuhan-Hu-1 strain (accession number NC_045512.2).

For Spike S2 protein amplification, the primers set used were: (i) First round (one step RT-PCR) (i) an outer primer pair COV_5_out_foward ACC AGG TTG CTG TTC TTT ATC AG (23379-23402pb) and COV_6_out_reverse ACTATGGCAATCAAGCCAGCT (25225-25246pb), producing a 1,844 kb fragment; followed for two nested PCR the primers used were (i) inner pair COV_5_out_foward ACCAGGTTGCTGTTCTTTATCAG (23379-23402pb) and COV_7_inner_Reverse GCACTTCAGCCTCAACTTTGT (24515-24536 pb), generating a fragment of 1,157kb (S2-A) and (ii) COV_8_out_foward GTGCAGGTGCTGCATTACA (24228–24247pb) and COV_6_out_reverse ACTATGGCAATCAAGCCAGCT (25225-25246pb) generating a fragment of 1,018kb each (S2-B). These pairs of primers permitted the partial amplification of the S2 protein (23545-25225 comprising to 561a.a), the complete S2 region comprising nucleotide position 23545 – 25371 (609 amino acid) based on SARS-CoV-2 Wuhan-Hu-1 strain (accession number NC_045512).

### Nucleic acid extraction

SARS-CoV-2 RNA was extracted from Nasopharyngeal (NP) swab samples by (QIAmp® viral RNA mini kit (Qiagen, Hilden, Germany; Biogene, Bioclean, Brazil) according manufacture’s protocol. Extraction followed the ongoing diagnosis routines at the laboratories and leftovers from this routines was kept in −70 °C until use.

### RT-PCR and nested PCR protocols

For both S1 and S2 Spike protein region, a similar one-step RT-PCR was designed. Extracted RNA was reverse-transcribed and amplified using SuperScript® III One-step RT-PCR system with Platinum Taq High Fidelity® (Life Technologies, USA). The total reaction mixture volume of 50μL contained the following: 2x reaction mix (25 μL), 10 pM primers (1μL each), enzyme mix (reverse transcriptase and Taq polymerase, 1μL), extracted viral RNA template (10μL), and RNase-free water (12μL). RT-PCR conditions for amplification were as follows: reverse transcription at 55°C for 30 min, initial PCR activation at 94°C for 2min, 35 amplification cycles of denaturation at 94°C for 30 s, annealing at 55°C for 30s, extension at 68°C for 2min 45s, and a final extension at 68°C for 10 min. For nested PCR, the RT-PCR product (2,5 µL), 10 pM primers (1 µL each), and RNase-free water (8 µL) were added to a Go Taq® Green Master Mix 2X (12,5 µL) (Promega Biosciences, CA). PCR conditions were as follows: initial denaturation at 94°C for 2 min, 35 cycles of denaturation at 94°C for 30s, annealing at 55°C for 30 s extension at 72°C for 2 min, and a final extension at 72°C for 10 min.

The products of RT-PCR and nested PCR were loaded in to a 1% agarose gel and visualized under ultraviolet light.

## Ethical approval

This study was carried out in accordance with the Declaration of Helsinki as revised in 2000, and approved by the Ethics Committee of the Adolfo Lutz Institute, São Paulo, Brazil. The study was registered at the institute, CTC 18M/2020 and CTC 39M/2020 and at the institutional ethical committee - CAAE: 31924420.8.0000.0059 and CAAE: 43250620.4.1001.0059.

All study participants were tested for SARS-CoV-2 at a public laboratory and has results made available to patients though GAL health system data. Those that did not provide informed consent had data anonymized prior to analysis and information used only for surveillance purposes.

### Sequencing

The 3,804 kb PCR product (partial Spike protein) amplification was sequenced using four primers for each region (Table1). Each sequencing reaction was performed using 4µL of BigDye Terminator v3.1 cycle sequencing kit® (Applied Biosystems) and 3,2µL for each primer (1µM) plus water to a final volume 20µL per reaction. Dye-labelled products were sequenced using a Genetic Analyzer ABI 3500 (Applied Biosystems). Sequencing chromatograms were edited manually using Sequencher 4.7 software (Gene Codes, USA). Sequences were analyzed in comparison to reference sequences but mutations list was generated at o CoVsurver tool for mutation analysis of hCoV-19 at GISAID (https://www.gisaid.org/epiflu-applications/covsurver-mutations-app/). Nucleotide sequences accession numbers are: EPI_ISL_1182103, EPI_ISL_1182104, EPI_ISL_1182101,EPI_ISL_1182102, EPI_ISL_1182099, EPI_ISL_1182100, EPI_ISL_1182097, EPI_ISL_1182098, EPI_ISL_1182095, EPI_ISL_1182096 EPI_ISL_1191781.

**Table 1.**
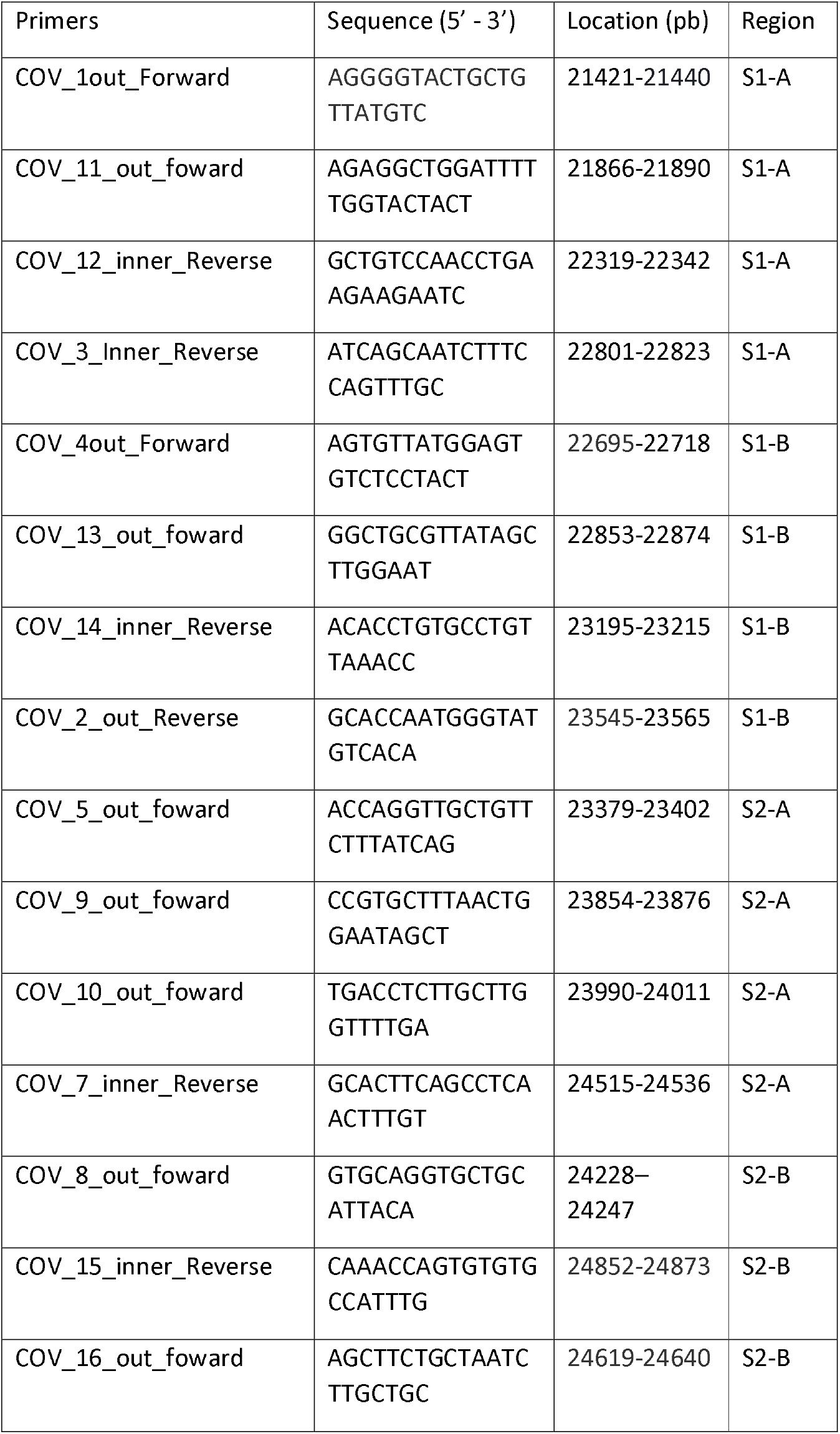
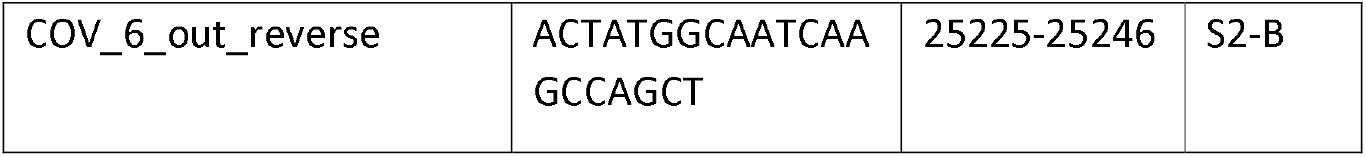
Sars-CoV-2 virus partial Spike (S1 and S2) region primer sets designed for Sanger sequencing

## Results

Table 2 depicts patient’s demographic data and respective cycle-threshold (CT) of rt-qPCR. All cases in this collected SWNF samples for COVID rt-qPCR test for diagnostic purposes. At the time of the study a symptomatic clinical settings was required for testing. Median time on symptoms was 4 (1-8) days.

**Table 2.** Demographic information of study patients. Table 2 shows age in years, gender (male or female), cycle threshold, CT, obtained for genes E and N, and city of sample collection, all at the State of Sao Paulo.

| **ID sequence** | **Age** | **Gender** | **Date of Collection** | **CT** | **Local of Collection** |
| --- | --- | --- | --- | --- | --- |
| hCov-19/Brazil/IAL-NDSS-02/21 | 58 | M | 17/01/2021 | 19/20 | Peruibe |
| hCov-19/Brazil/IAL-NDSS-03/21 | 19 | M | 14/01/2021 | 28/28 | Cubatao |
| hCov-19/Brazil/IAL-NDSS-04/20 | 44 | F | 04/09/2020 | 26/24 | Itaquaquecetuba |
| hCov-19/Brazil/IAL-NDSS-05/20 | 70 | F | 01/06/2020 | 17/18 | Sao Jose dos Campos |
| hCov-19/Brazil/IAL-NDSS-05/21 | 43 | F | 17/01/2021 | 17/17 | Praia Grande |
| hCov-19/Brazil/IAL-NDSS-06/21 | 22 | F | 16/01/2021 | 20/21 | Praia Grande |
| hCov-19/Brazil/IAL-NDSS-07/21 | 68 | F | 17/01/2021 | 29/28 | Praia Grande |
| hCov-19/Brazil/IAL-NDSS-09/21 | 35 | F | 17/01/2021 | 19/20 | Praia Grande |
| hCov-19/Brazil/IAL-NDSS-10/21 | 44 | F | 17/01/2021 | 19/17 | Praia Grande |
| hCov-19/Brazil/IAL-NDSS-11/21 | 44 | F | 19/04/2020 | 18/19 | Cubatao |
| hCov-19/Brazil/IAL-NDSS-12/21 | 34 | F | 16/04/2020 | 23/23 | Sao Vicente |

## Sequence results

All the 11 sequences had the D614G amino acid change compared to Wuhan reference sequence, but 82% 9/11 had two or more additional changes. Table 3 depicts sequence and list of mutations identified at Spike protein. Key mutations characteristics of the P1 variant were identified in two sequences of 2021 (samples hCov-19/Brazil/IAL-NDSS-05/21 and hCov-19/Brazil/IAL-NDSS-07/21). Some mutations, as E484K, were frequent, present in six sequences of this small data set, two with the P1 amino acid mutation complex and in 4 other genomes. The presence of E484K was observed in 6 out of 7 sequences of 2021, but not (0/4) in those of 2020 (p=0.015, Fisher Exact, two-tailed). Table 3 summarizes some information on the sequences obtained in this study as fragment length of the sequences in amino acids and nucleotides with your respective mutations and clade according to GISAID. Sequences may be accessed at GISAID, accession numbers are: EPI_ISL_1182103, EPI_ISL_1182104, EPI_ISL_1182101, EPI_ISL_1182102, EPI_ISL_1182099, EPI_ISL_1182100, EPI_ISL_1182097, EPI_ISL_1182098, EPI_ISL_1182095, EPI_ISL_1182096 EPI_ISL_1191781.

**Table 3.** sequence identification, lenght and mutations of the patients samples.

| ID sequence | Fragment Length(nt) | Fragment Length(aa) | Fragment Range | Mutations | Clade |
| --- | --- | --- | --- | --- | --- |
| hCov-19/Brazil/IAL-NDSS-02/21 | 2551 | 849 | 22695nt - 25245nt | D614G, V1176F, I1225L | G |
| hCov-19/Brazil/IAL-NDSS-03/21 | 3785 | 1257 | 21461nt - 25245nt | E484K, D614G, V1176F, I1225L | G |
| hCov-19/Brazil/IAL-NDSS-04/20 | 2862 | 953 | 22384nt - 25245nt | D614G, V1176F, I1225L | G |
| hCov-19/Brazil/IAL-NDSS-05/20 | 2845 | 945 | 22402nt - 25245nt | E281K, D614G, T1027I, V1176F | G |
| hCov-19/Brazil/IAL-NDSS-05/21 | 3785 | 1257 | 21461nt - 25245nt | L18F, T20N, P26S, G72V, D138Y, R190S, K417T, E484K, N501Y, D614G, H655Y, T1027I, V1176F | G |
| hCov-19/Brazil/IAL-NDSS-06/21 | 2840 | 946 | 22406nt - 25245nt | E484K, D614G, V1176F | G |
| hCov-19/Brazil/IAL-NDSS-07/21 | 2095 | 694 | 21461nt - 23555nt | L18F, T20N, P26S, D138Y, R190S, L229F, K417T, E484K, N501Y, D614G, A653P, H655Y | G |
| hCov-19/Brazil/IAL-NDSS-09/21 | 2730 | 909 | 22517nt - 25245nt | E484K, D614G, A623S, V1176F, I1225L | G |
| hCov-19/Brazil/IAL-NDSS-10/21 | 2832 | 943 | 22404nt - 25235nt | E484K, D614G, V1176F | G |
| hCov-19/Brazil/IAL-NDSS-11/21 | 3066 | 1018 | 21460nt - 24525nt | D614G | G |
| hCov-19/Brazil/IAL-NDSS-12/21 | 2048 | 678 | 21461nt - 23508nt | D614G | G |

## Discussion

In this small study we describe preliminary data from an ongoing effort to develop simple RT-PCR protocols for obtaining Spike region sequences covering key amino acid mutations associated to the most concerning variants recently identified. The protocol was able to generate good quality sequences that may cover most (up to 95%) of the spike gene. This protocol does not yet cover all Spike protein, but additional segments of amplified DNA will allow full Spike coverage. The protocol however, can provide the identification of the key amino acid mutations that have been associated to increase transmissibility, covers most regions that have been associated to viral pathogenesis, as the RBD and may identify the UK, SA and BR variants. Surveillance initiatives may use these key amino-acid mutations at positions that are present in the three major variants to allow the identification these variants based in the Spike signatures. Selected samples may be further evaluated with NGS.

We found a high proportion of E484K mutation at Spike protein, 6 out of these 11 sequences harbor the mutation, only two of that with the other additional amino acid mutations that are characteristics of P1 variant. This E484K change may affect recognition of host cells and may “weakens the potency of antibodies that can ordinarily disable the virus” (Ewen 2021). In vitro studies with a new B.1.1.7 carrying the E484K mutation increases the amount of serum antibody needed to block cell infection (Collier 2021).

The fact that the other P1 mutations are not present among with E484K in many samples may suggest that new variants may be evolving independently in the region that carries mutations useful for the viral life cycle, as due to immune scape potential and/or longer binding to cell receptors.

We opt to release this preliminary data to stimulate other groups that may benefit from the simplicity and potential informative power of a simpler approach to genomic surveillance. If in one hand the methodology of partial sequencing does not allow proper lineage evaluation, it can provide a swift access to information on the presence of these key mutations among the samples of a region or a subgroup of the population. If linked to proper contact tracing and preventive measures, it may provide a powerful tool to block variant expansion to new areas and populations. Therefore, this approach may be an alternative and sum to build the necessary increase in genomic surveillance. The NGS capability is currently limited, especially but not only in resource-limited settings. In Brazil, for example, with over 11 million document cases, fewer than 3,000 sequences registered at sequence databanks.

Shared cycling temperatures used in these protocols may optimized thermal cycler equipment use, and adjustments protocols are being tested to reduce cost. Small adjustments in sample processing to better adapt to real world situations may improve surveillance capability. We are currently testing simpler alternative protocols that may be easier to be implemented in resource-limited settings to improve surveillance capability. This and alternative approaches needed to be further tested for feasibility as the idea of simpler protocols to contribute to the monitoring of SAR-CoV-2 evolution may prove valuable.

## Data Availability

Sequence deposite at GISAID, non confidential data not already submitted with sequences available upon request

